# Reduction in preterm births during the COVID-19 lockdown in Ireland: a natural experiment allowing analysis of data from the prior two decades

**DOI:** 10.1101/2020.06.03.20121442

**Authors:** RK Philip, H Purtill, E Reidy, M Daly, M Imcha, D McGrath, NH O’Connell, CP Dunne

## Abstract

**Background:** Aetiology of preterm birth (PTB) is heterogeneous and preventive strategies remain elusive. Socio-environmental measures implemented as Ireland’s prudent response to the SARS-CoV-2 virus (COVID-19) pandemic represented, in effect, a national lockdown and have possibly influenced the health and wellbeing of pregnant women and unborn infants. Cumulative impact of such socio-environmental factors operating contemporaneously on PTB has never been assessed before.

**Methods:** Regional PTB trends of very low birth weight (VLBW) infants in one designated health area of Ireland over two decades were analysed. Poisson regression and rate ratio analyses with 95% CI were conducted. Observed regional data from January – April 2020 were compared to historical regional and national data and forecasted national figures for 2020.

**Results:** Poisson regression analysis found that the regional historical VLBW rate per 1000 live births for January to April, 2001-2019 was 8.18 (95% CI: 7.21, 9.29). During January to April 2020, an unusually low VLBW rate of just 2.17 per 1000 live births was observed. The rate ratio of 3.77 (95% CI: 1.21, 11.75), p = 0.022, estimates that for the last two decades there was, on average, 3.77 times the rate of VLBW, compared to the period January to April 2020 during which there is a 73% reduction. National Irish VLBW rate for 2020 is forecasted to be reduced to 400 per 60,000 births compared to the historical 500-600 range.

**Conclusion:** An unprecedented reduction in PTB of VLBW infants was observed in one health region of Ireland during the COVID-19 lockdown. Potential determinants of this unique temporal trend reside in the summative socio-environmental impact of the COVID-19 dictated lockdown. Our findings, if mirrored in other regions that have adopted similar measures to combat the pandemic, demonstrate the potential to evaluate these implicated interdependent behavioural and socio-environmental modifiers to positively influence PTB rates globally.

**Key Questions:** *What is already known?:*

- Premature birth is an important contributor for under-five mortality globally.
- Currently there is no broadly accepted and effective strategy to prevent the birth of premature very low birth weight infants.
- Impact of socio-environmental and maternal behavioural modifications on the incidence of preterm birth has not been assessed.

*What are the new findings?:*

- COVID-19-triggered national lockdown in Ireland created an opportunity to study the cumulative influence of socio-environmental modifications on pregnant mothers.
- An unprecedented 73% reduction in the rate of very low birth weight deliveries was noted in one designated health region of Ireland during January to April of 2020 in comparison to the preceding 20 year timeframe.
- Our observations, if nationally mirrored, indicate that birth rate of very low birth weight premature infants in Ireland is forecasted to decrease considerably in 2020.

*What do the new findings imply?:*

- Socially rooted modifiers such as family support, work related stress and commuting, environmental pollution, infection avoidance, sleep and nutritional support, adequate exercise, reduced exposure to tobacco and illicit drugs, avoidance of financial strain, all cumulatively could contribute to reduce preterm birth rate.
- Our observations, if reflected in other countries that adopted COVID-19-prompted lockdown measures, would redefine the antecedents that trigger the yet poorly understood pathways leading to preterm births.
- Prematurity rate would be the most important ‘*curve to bend*’ in the context of reducing infant mortality globally and thus promote the achievement of sustainable development goals for children.

## Introduction

Over 15 million babies are born too early, too sick and too small in the world every year. One million of these infants die.^1^ Preterm birth (PTB), below 37 weeks of gestation, contributes significantly to infant mortality.^2,3,4,5^ In 2016, 46% of the under-five mortality globally was contributed to by neonatal deaths within the first 28 days of life and the main contributor was prematurity.^5^ PTB rate is increasing in most developed and developing countries and a significant proportion of spontaneous PTB is of unknown aetiology.^2,6^ The frequency of PTB varies from 5-9% in Europe, 10.6% in North America to 11.9% in Africa.^7,8^ While repeated PTB is reported, most preterm very low birth weight (VLBW, <1,500 gm) and extremely low birth weight (ELBW, <1,000 gm) infants are born to women with no prior history of PTB.^5,9^

Prevention of PTB is considered a public health priority. Despite the obvious and growing relevance, progress has been modest.^10^ Currently there is no standardised and effective strategy to prevent PTB, and implementation of socio-cultural approaches to mitigate the risk of PTB would require better elucidation of non-medical factors that are both under-recognised and under-evaluated.^11^ Although the causal biological mechanisms mediating PTB are poorly understood, preterm premature rupture of membranes (PPROM) complicates one third of all PTB.^12^ Suggested alternate biological antecedents include amnionic inflammasome, alterations to vaginal microbiota, variations in cytokines, chemokines and other inflammatory modulators, as well as intra-amniotic inflammation (IAI) and infections.^13,14,15^ It has been suggested that socio-environmental phenotype could offer insights and recognising the sources of heterogeneity and phenotypic plasticity in PTB may inform eventual effectiveness of preventive measures.^16,17^

Ireland offers a unique opportunity to evaluate PTB as it has been one of the very few developed countries that legally prohibited termination of pregnancy (TOP) or abortion until the ‘*Regulation of Termination of Pregnancy Act*’ was passed on 20^th^ December 2018 with services initiated on 1^st^ January 2019.^18^ In that specific setting, there existed an opportunity to compare data from the pre-TOP era with 2020 data to determine the potential influence on PTB rates due to socio-environmental measures implemented as Ireland’s prudent response to the SARS-CoV-2 virus (COVID-19) pandemic. In effect, these measures constituted a national lockdown and may possibly have influenced the health and wellbeing of pregnant women and their unborn infants. We wished to assess these redefined social and behavioural boundaries that could foster an environment encompassing elements that are favourable to pregnancies reaching full term.

## Methods

### Setting

A nationwide lockdown was adopted in Ireland on 12^th^ March 2020 in response to the COVID-19 pandemic and was extended to 18^th^ of May with the shutdown of offices, shops, colleges, schools, childcare facilities and all other institutions deemed non-essential. Traffic and mobility restrictions were imposed and most of the workforce had to adapt to a new work-from-home (WFH) model. Department of Health, National Public Health Emergency Team (NPHET) and Health Service Executive (HSE) of Ireland were advising the population from mid-February 2020 to follow strict hand hygiene measures, social distancing and adherence to WHO recommendations to reduce COVID-19 transmission. This period of national lockdown and the pre-lockdown weeks of extra healthcare vigilance and restrictions resulted in holistic alterations to the wellbeing of pregnant women. Potential influence of a multitude of biological, physical and environmental factors could cumulatively influence and modify the PTB of VLBW infants. These few months offered a unique opportunity to study the effects of a *‘Nature’s experiment’* of non-medical, behavioural and socio-environmental alterations as the key determinants restoring overall health of the ‘intrauterine habitat’ and influencing the continuation of foetal life.

### Study population

University Maternity Hospital Limerick (UMHL), serving as the only maternity facility to a population of 473,000 from the counties of Limerick, Clare, North Tipperary and nearby catchment areas provides a unique opportunity to analyse the demographic and epidemiologic trends of births involving VLBW and ELBW infants of one of the designated health regions in the Republic of Ireland.^19^ All preterm infants from 22 weeks of gestation are treated locally (apart from surgical or cardiac interventions) and our perinatal demography and patient characteristics of the very preterm population have already been published.^20^ PTB involving VLBW infants from 22 weeks of gestation onwards at UMHL from 1^st^ January 2001 to 30^th^ April 2020 were included in the study. Retrospective descriptive datasets were linked from the labour ward register, neonatal admission register and pre-submission data towards the Vermont Oxford Network (VON) international benchmarking.^21^ No cases were excluded based on congenital anomalies, multiple gestations or inconsistencies around gestational age estimation. We did not sub-classify PTB to spontaneous onset of labour with intact membranes, PPROM or medically initiated labour onset through induction or caesarean.^7^ Our VLBW trends were compared with the published National figures from Central Statistics Office (CSO) and the National Perinatal Epidemiology Centre (NPEC) of Ireland.^22,23^

### Hypothesis

There were no overarching significant alterations to the antenatal, obstetric or intrapartum care pathways initiated for the pregnant women at UMHL or our health region from January-April of 2020 compared to preceding years. This offered us the opportunity to hypothesise whether the non-medical, community-based, socio-environmental determinants and modifiable behavioural factors brought on by the Irish public health responses to COVID-19 pandemic and lockdown would be of significance to our observed temporal trends in PTB rates.

### Statistical analysis

Fully anonymised and de-identified dataset fulfilling general data protection regulation (GDPR) compliance was prepared for statistical analysis.^24^ Significance of temporal trends in the VLBW and ELBW rates per 1000 births were assessed using Poisson regression, where time was entered as a continuous variable. Poisson regression with 95% Wald confidence intervals and rate ratio analysis were used to compare the observed VLBW rate for Jan-April of 2020 at UMHL to historical data. Estimates of the prevalence of VLBW and ELBW per 1000 births pre-2020 assessed the potential impact of January – April 2020 regional data on the national expectation of VLBW for 2020 in Ireland (based on the previous published Irish data from VON and CSO).^21,22,23^ All data were analysed using IBM SPSS Statistics V.26.

### Patient and Public Involvement

Patient and public involvement (PPI) was initiated at the outset of research planning. Irish Neonatal Health Alliance (INHA), the patient advocacy group (Registered Charity Number: 20100100) representing parents of newborn infants in Ireland and member of European Foundation for the Care of Newborn Infants (EFCNI), was invited at the design stage of the study and was a signatory in the Research Ethics submission. INHA reviewed the aims of the study and confirmed that the issues addressed through the research is of relevance to patients and public. INHA reaffirmed that the patient confidentiality is not breached at any stage of the research, and specifically requested forecasting of National VLBW births through appropriate analysis in order for the advocacy group to prioritise their family centred care programmes for 2020. INHA also nominated a parent of a premature VLBW infant to be an independent external reviewer of VLBW and ELBW data for 2020. Designated representative of the patient advocacy group is a member of the study team and authorship of the manuscript. Once published, the relevant findings of the study will be disseminated through the websites of INHA www.inha.ie and EFCNI www.efcni.org Guidance for Reporting Involvement of Patients and the Public (GRIPP2) reporting checklist as applicable to this study has been fulfilled. (Appendix 1)

### Data verification and Reporting

Considering the unprecedented and significant reduction of the VLBW and ELBW numbers observed during the lockdown and pre-lockdown phases of extra public health vigilance, we have verified the accuracy and authenticity of primary data capture with external independent professionals and patient representative (as part of the PPI initiative) who are not members of the research team or authorship.

### Ethics Approval

University Hospital Limerick Research Ethics Committee approval was granted for the study.

## Results

Over the last twenty years UMHL had 93,018 live births and during the four months of January to April from 2001 to 2020 there were 30,705 live births. Annual live births, annual ELBW and VLBW rates as well as the respective numbers for 1^st^ January to 30^th^ April of each year for the last two decades are summarised in Table 1. Poisson regression analyses of the 2001 – 2019 data did not find any evidence of temporal trends for UMHL January – April VLBW (Wald Chi-square= 0.784, p = 0.376), January – April ELBW (Wald Chi-square = 0.464, p = 0.496), annual VLBW (Wald Chi-square = 0.366, p = 0.545) or annual ELBW (Wald Chi-square = 0.008, p = 0.929).

**Table 1:** University Maternity Hospital Limerick (UMHL) Regional data of VLBW and ELBW births for 2001-2020.

| January - April |  |  |  |  |  | Annual |  |  |  |  |
| --- | --- | --- | --- | --- | --- | --- | --- | --- | --- | --- |
| Year | Live Births | ELBW |  | VLBW |  | Live Births | ELBW |  | VLBW |  |
|  |  | Cou<br>nt | Rate/10<br>00 | Cou<br>nt | Rate/10<br>00 |  | Cou<br>nt | Rate/10<br>00 | Cou<br>nt | Rate/10<br>00 |
| **2001** | 1337 | 4 | 2.99 | 12 | 8.98 | 4042 | 15 | 3.71 | 32 | 7.92 |
| **2002** | 1428 | 2 | 1.40 | 15 | 10.50 | 4371 | 6 | 1.37 | 31 | 7.09 |
| **2003** | 1498 | 5 | 3.34 | 13 | 8.68 | 4514 | 16 | 3.54 | 42 | 9.30 |
| **2004** | 1458 | 7 | 4.80 | 12 | 8.23 | 4418 | 20 | 4.53 | 37 | 8.37 |
| **2005** | 1447 | 3 | 2.07 | 13 | 8.98 | 4411 | 10 | 2.27 | 41 | 9.29 |
| **2006** | 1539 | 4 | 2.60 | 12 | 7.80 | 4692 | 16 | 3.41 | 47 | 10.02 |
| **2007** | 1704 | 6 | 3.52 | 18 | 10.56 | 5153 | 12 | 2.33 | 55 | 10.67 |
| **2008** | 1818 | 4 | 2.20 | 12 | 6.60 | 5443 | 12 | 2.20 | 38 | 6.98 |
| **2009** | 1803 | 6 | 3.33 | 17 | 9.43 | 5432 | 13 | 2.39 | 44 | 8.10 |
| **2010** | 1676 | 6 | 3.58 | 14 | 8.35 | 5233 | 17 | 3.25 | 52 | 9.94 |
| **2011** | 1671 | 4 | 2.39 | 10 | 5.98 | 5137 | 11 | 2.14 | 27 | 5.26 |
| **2012** | 1655 | 6 | 3.63 | 12 | 7.25 | 4905 | 17 | 3.47 | 43 | 8.77 |
| **2013** | 1580 | 2 | 1.27 | 11 | 6.96 | 4594 | 13 | 2.83 | 42 | 9.14 |
| **2014** | 1482 | 3 | 2.02 | 10 | 6.75 | 4522 | 13 | 2.87 | 36 | 7.96 |
| **2015** | 1565 | 4 | 2.56 | 10 | 6.39 | 4690 | 12 | 2.56 | 45 | 9.59 |
| **2016** | 1483 | 4 | 2.70 | 14 | 9.44 | 4473 | 10 | 2.24 | 34 | 7.60 |
| **2017** | 1406 | 7 | 4.98 | 14 | 9.96 | 4416 | 16 | 3.62 | 35 | 7.93 |
| **2018** | 1464 | 3 | 2.05 | 9 | 6.15 | 4439 | 11 | 2.48 | 37 | 8.34 |
| **2019** | 1310 | 8 | 6.11 | 12 | 9.16 | 4144 | 15 | 3.62 | 31 | 7.48 |
| **2020** | 1381 | 0 | 0.00 | 3 | 2.17 |  |  |  |  |  |

The VLBW rate per 1000 live births (January – April) based on aggregate 2001 – 2019 data was 8.18 (95% Wald CI: 7.21, 9.29), representing a forecast January – April 2020, in the absence of a temporal trend. However, the observed total number of only three VLBW in January – April 2020 (all of them born before mid-February 2020, with no ELBW) confirms an unusually low VLBW rate of just 2.17 per 1000 live births (95% Wald CI: 0.70, 6.74), 73% lower than the forecasted rate of 8.18. (Figure 1). The Rate Ratio comparing the risk of VLBW in January – April 2001-2019 to that from January – April in 2020 was 3.77 (95% Wald CI: 1.21, 11.75), p = 0.022, suggesting that for two decades pre-2020 there was 3.77 times the rate of VLBW for the months of January – April in comparison to 2020. The ELBW rate per 1000 live births for January – April using aggregated data from 2001 – 2019 was 3.0 (95% Wald CI: 2.43, 3.70). There were no ELBW live births recorded for the health region during January – April 2020. (Figure 2)

**Figure 1:**
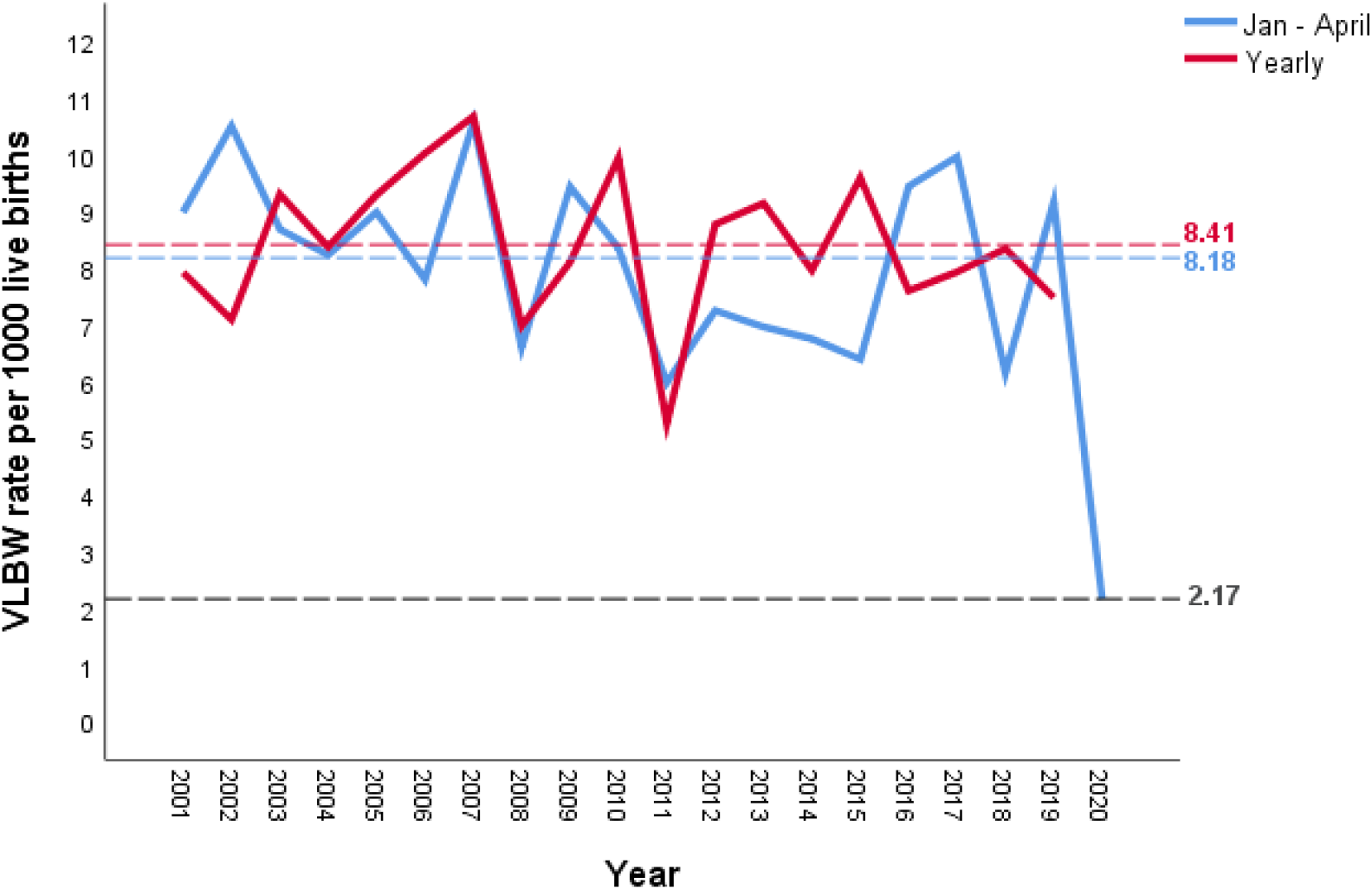
VLBW births from January – April and Yearly for UMHL from 2001-2020.

**Figure 2:**
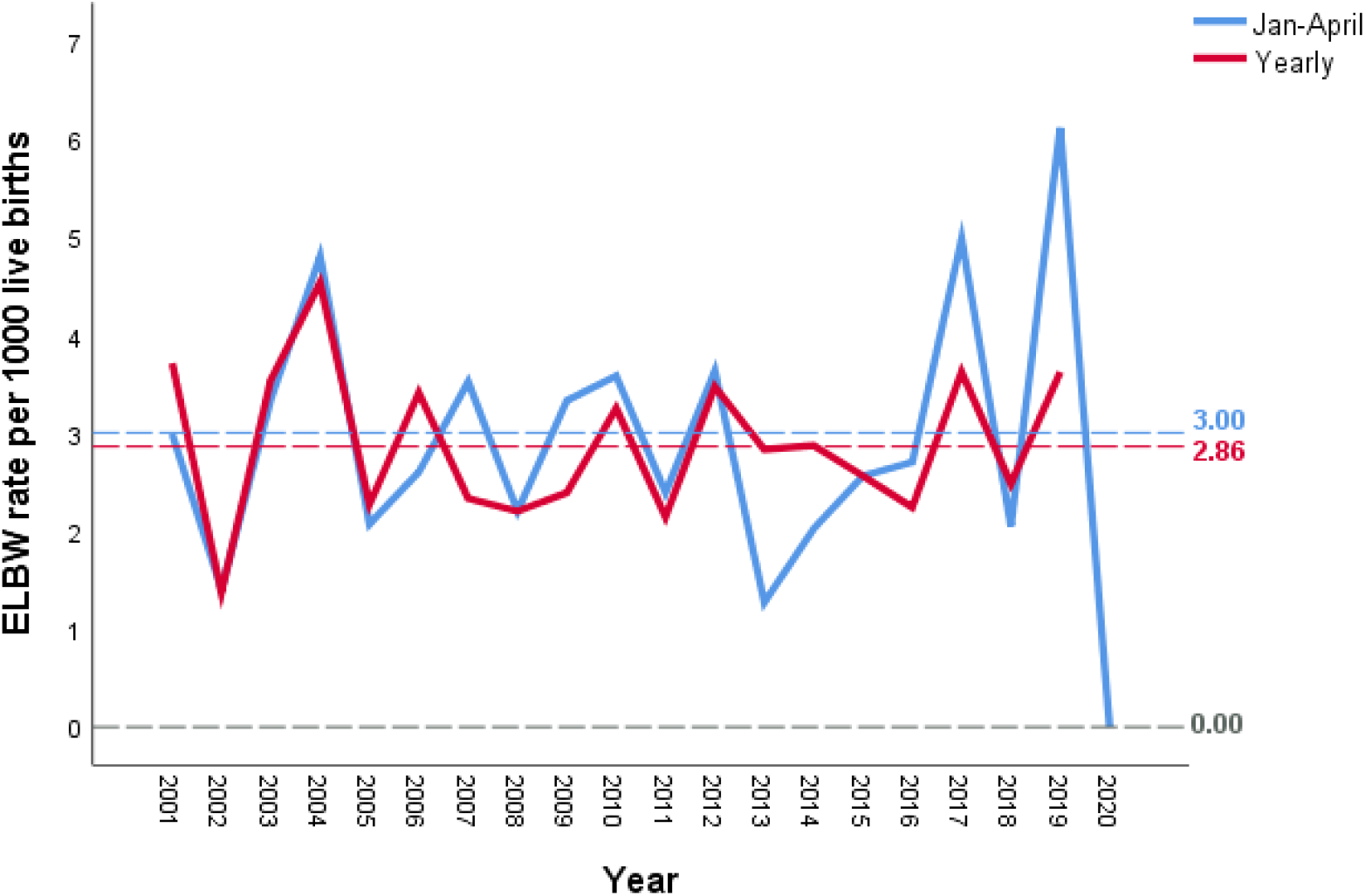
ELBW births from January – April and Yearly for UMHL from 2001-2020.

The VLBW rate per 1000 live births from the aggregated regional numbers for 2001 - 2019 yearly data was computed to be 8.41 (95% Wald CI: 7.83, 9.04), providing a forecasted annual rate for 2020. A continuation of the 73% reduction from forecast in the January-April VLBW rate for the health region for the remainder of 2020 would forecast a rate of 2.27 VLBW per 1000 live births. However, a reversal to the regional population’s pre-lockdown behavioural and socio-environmental *status quo* could increase the rate towards the historical rates.

The forecast for the 2020 national VLBW rate is taken from the 2014-2017 VON date for Ireland, giving a mean of 9.37 per 1000 live births, no trend is evident. (Table 3) If the national VLBW rate reflects the observed regional rate of 2.17 (95% Wald CI: 0.70, 6.74) (January-April 2020), the upper confidence limit estimates the number of VLBW infants to be born in Ireland in 2020 could be reduced to approximately 400 per 60,000 births (historically 500-600). (Table 2) However, variation in the population’s compliance with lockdown in other regions of the country and post-lockdown deterioration in behavioural and socio-environmental factors or the possibility of a ‘baby boom’ in late 2020 could regain the PTB rate back to the historical over 500 per 60,000 births.

**Table 2:**
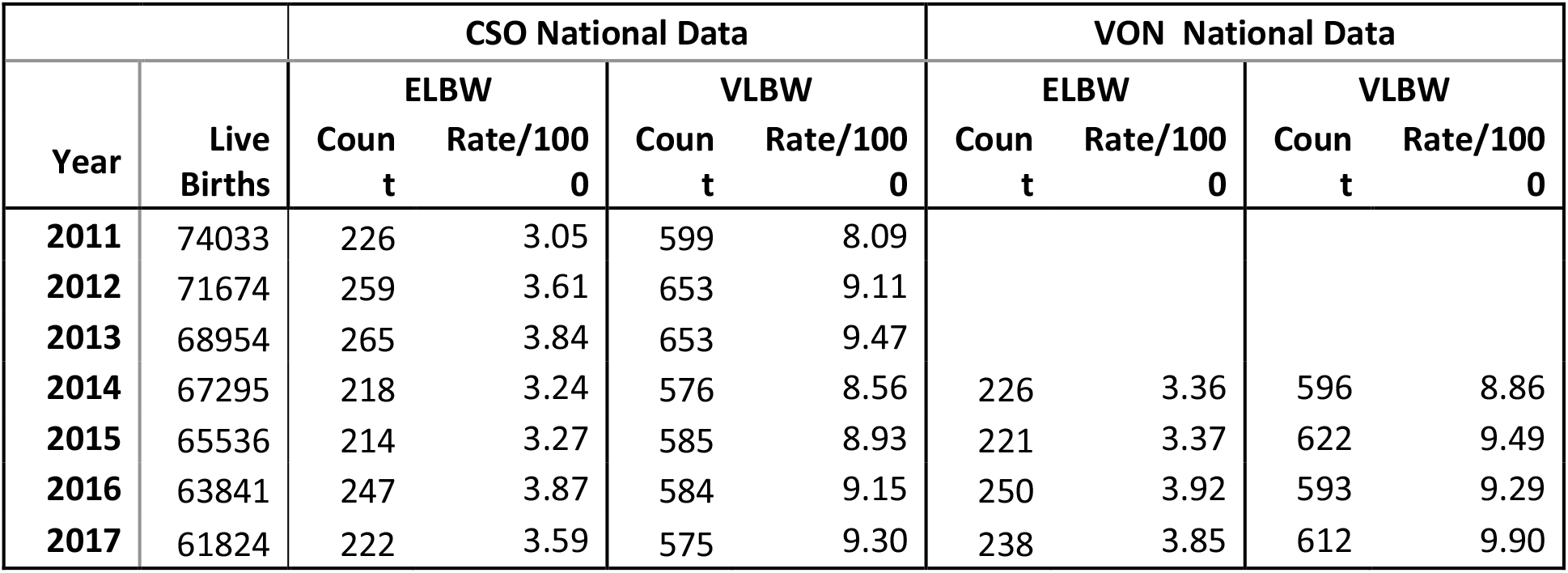
National Irish data for VLBW and ELBW births based on the published CSO and VON numbers. CSO-Central statistics office. VON-Vermont Oxford Network.

|  |  | CSO National Data |  |  |  | VON National Data |  |  |  |
| --- | --- | --- | --- | --- | --- | --- | --- | --- | --- |
| Year | Live Births | ELBW |  | VLBW |  | ELBW |  | VLBW |  |
|  |  | Coun<br>t | Rate/100<br>0 | Coun<br>t | Rate/100<br>0 | Coun<br>t | Rate/100<br>0 | Coun<br>t | Rate/100<br>0 |
| 2011 | 74033 | 226 | 3.05 | 599 | 8.09 |  |  |  |  |
| 2012 | 71674 | 259 | 3.61 | 653 | 9.11 |  |  |  |  |
| 2013 | 68954 | 265 | 3.84 | 653 | 9.47 |  |  |  |  |
| 2014 | 67295 | 218 | 3.24 | 576 | 8.56 | 226 | 3.36 | 596 | 8.86 |
| 2015 | 65536 | 214 | 3.27 | 585 | 8.93 | 221 | 3.37 | 622 | 9.49 |
| 2016 | 63841 | 247 | 3.87 | 584 | 9.15 | 250 | 3.92 | 593 | 9.29 |
| 2017 | 61824 | 222 | 3.59 | 575 | 9.30 | 238 | 3.85 | 612 | 9.90 |

## Discussion

Prematurity poses significant medical, emotional, physical, psychological and financial burden for the affected infants, their support network, health systems, economies and society as a whole.^25^ PTB through significant neonatal morbidity also leads to long-term health concerns during childhood.^26^ A myriad of etiologic and antecedent factors could trigger PTB and the effectiveness of preventive measures depends on our precise understanding of the causation. A pan-European study found rising PTB rates in most countries and an increase in multiple births as well as assisted reproduction techniques (ART) also contributed to the overall increase. Understanding cross-country differences also could inform strategies for PTB reduction.^27^

Pregnancy is an ideal opportunity to encourage positive behavioural changes.^28^ Pregnancy Risk Assessment Monitoring System (PRAMS) in Ireland, National Institute of Clinical Evidence (NICE) guidelines as well as the *‘Safer Maternity Care’* document in UK are worthy initiatives aimed at PTB reduction.^28,29,30^ However, the yet under-recognised, behavioural, socio-cultural and socio-environmental modifications and opportunities designed to prolong the intrauterine nurturing milieu could offer far more to reduce PTB rates.

Prenatal period and foetal growth could be regarded as a matrix for our lives and societies.^31^ PTB also could have an evolutionary basis of predictive adaptive response congruous with the Developmental Origins of Health and Disease (DOHaD).^32,33^ Heterogeneous origins of PTB rate could be influenced by environmental changes, modifiable population factors, nutritional variations, stress factors and socioeconomic status.^17,33^

### Potential influence of the termination of pregnancy and stillbirths

As one of the very few developed regions of the world with abortion legally banned till late 2018, Ireland offers a unique opportunity to evaluate the natural history of PTB and the wider relation to socio-environmental alterations. Could the very low ELBW and VLBW figures in early 2020 be explained partially by the change in TOP law, allowing for TOP beyond 12 weeks gestation in the presence of major congenital anomalies (MCA) that limits foetal or neonatal viability? Examination of our regional and national historical data suggest not.

Historical mean prevalence of MCA among our regional cohort of VLBW from 2000 to 2018 (two decades of no TOP) was a mean of 9.2 % and the national mean for 2014, 2015, 2016 and 2017 were 9%, 7%, 9% and 8% respectively (55/596, 42/622, 54/593 and 51/612).^23^ That is, less than 1 in every 10 VLBW had the presence of MCA when no TOP was available. The TOPs undertaken in 2019 and 2020 from January-April (two in 2019 and four in 2020) beyond 12 weeks of gestation were for MCA in our region, in accordance with the national guidelines on compassionate grounds.^18^ There was no increasing trend for the stillbirth rate (SBR) of UMHL or the region during the study period, confirming no ‘displacement of vital statistics’ as an explanation for the VLBW reduction.

### COVID-19 lockdown triggered socio-environmental and behavioural modifiers

The *‘Nature’s experiment’* through the COVID-19 lockdown triggered unparalleled and widespread socio-environmental alterations which we hypothesise as the plausible explanation for PTB of VLBW rate to fall from 8.18 (95% CI: 7.21, 9.29) to 2.17 (95% CI: 0.70, 6.74) per 1000 live births for the January – April period. Specific modifiers, both facilitators and barriers, in the socio-cultural and socio-environmental settings that in our view would have influenced the mother-foetus pair to reduce the chances for PTB during the COVID-19 lockdown and pre-lockdown weeks of enhanced public health vigilance are summarised below (Figure 3).

**Figure 3:**
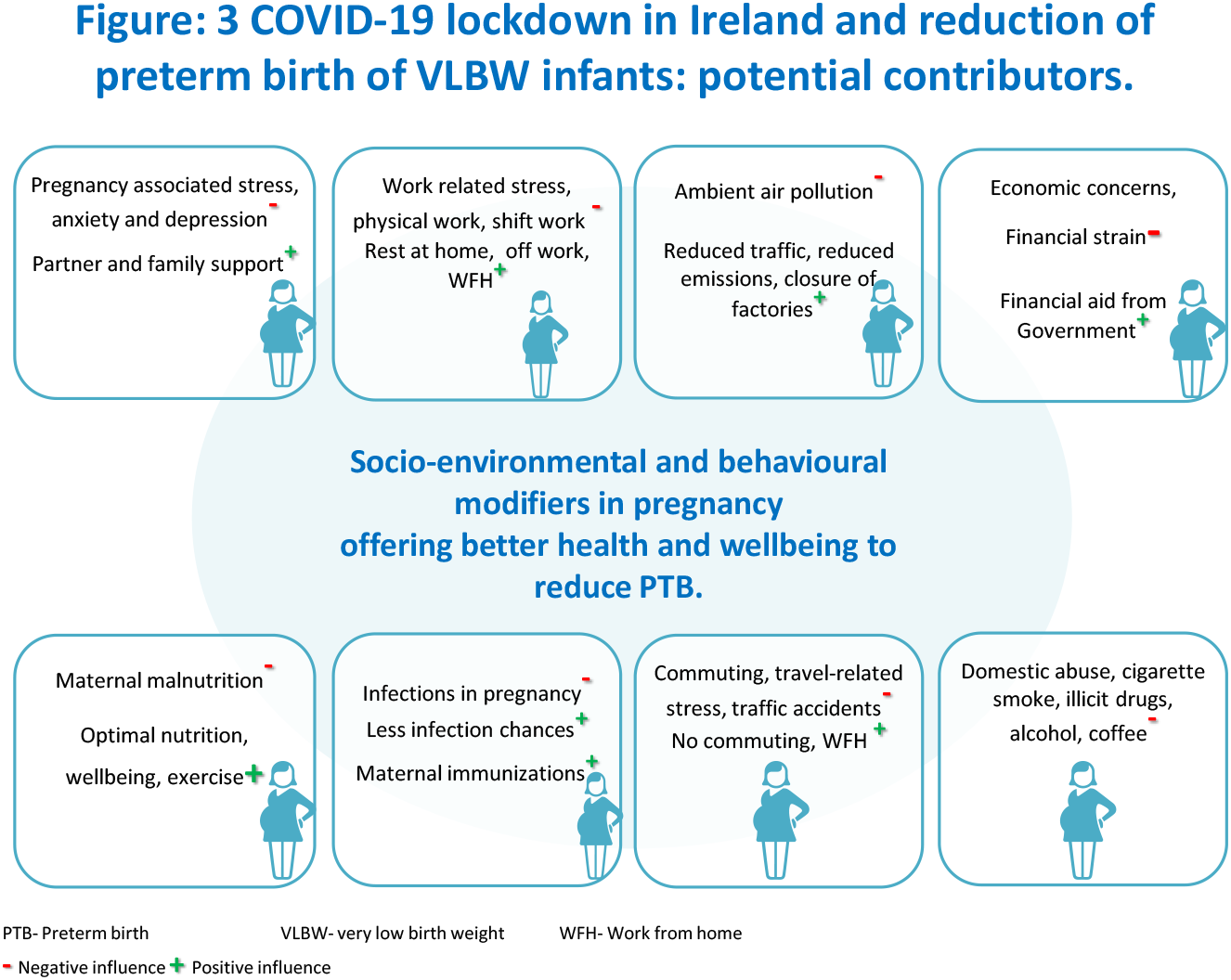
How the COVID-19 lockdown in Ireland possibly contributed to the reduction of preterm birth of VLBW infants.

#### 1. Pregnancy associated stress, anxiety and support systems

Psychological stresses increase the likelihood of PTB. Positive benefits from familial support have been reported.^9^ Male partner involvement on reducing PTB rates has been suggested.^34^ Compared to women who reported no partner involvement and support during pregnancy, those who had it reported better psychological well-being.^35^ There is strong evidence that pregnancy-specific anxiety, depression and stress increase the likelihood PTB.^36^ Impacts of on-call work during pregnancy in relation to stress and sleep are not well established.^37^ Measures of psychological distress including alterations in blood corticotrophin-releasing hormone (CRH) levels and cortisol are associated with PTB.^38^ The biologic pathways underlying stress-induced PTB remain poorly understood.^39^ We postulate therefore that the reduced pregnancy associated stress and increased support systems available during the COVID-19 lockdown could be contributory factors in the low levels for PTB of VLBW seen in the region.

#### 2. Work related stress, physical work, shift work and on-calls

Studies suggest an association of shift work, fixed night shifts and long working hours during pregnancy with PTB.^40,41^ Women working more than 55.5 hours *(vs* 40 hours) per week had a 10% increase in the odds of having a PTB.^40^ Danish National birth cohort study associated shift work with an increase in small-for-gestational-age (SGA).^42^

Physically demanding work while pregnant increases PTB as per systematic review and meta-analysis.^40,43^ A threefold increase in PTB was reported in women whose daily work entailed trunk bending for over one hour.^44^ Bed rest in hospital or at home is widely recommended as a conventional practice to prevent PTB through the possible reduction of uterine activity. While sufficient rest and relaxation seems appropriate, Cochrane review concluded neither supporting nor refuting the use of prolonged bed rest at home or in hospital, to prevent PTB.^45^ It is possible that the change in work practices due to the COVID-19 lockdown had the added benefit of reducing prematurity.

#### 3. Environmental factors, temporal variations, air pollution

A Unites States study exploring temporal patterns of PTB observed a periodicity for PTB rates.^46^ Association between ambient air pollution (AAP) exposure during pregnancy and PTB has been reported.^26,47,48^ Particulate matter, nitrogen dioxide, ozone, and carbon monoxide were the most commonly used markers of AAP.^47,48^ Sulphur dioxide (SO_2_) was the largest contributor to increase PTB and other agents were particulate matter (PM)_2.5 micron_, PM_10 micron_ and NO_2_.^26^ One study from Wuhan, China linked PTB to the atmospheric pollutant, vanadium.^49^ The relationship between atmospheric temperature and seasonal fluctuations to PTB has been suggested. European pattern is discerned with a spring peak and in Asia and US a seasonal variance of up to 5-10% has been reported.^50^

No major meteorological events were reported in Ireland during the 2020 lockdown in comparison to the preceding two decades and the observed decline in our VLBW rate is well beyond the variations in the natural annual temporal trends reported to date. Cumulative contribution to AAP improvement by a reduction in all modes of transport, reduced consumption of diesel and petrol, reduced production and distribution of goods and closure of factories, all led to reduced environmental pollution in UK and Europe during the lockdown.^51^

#### 4. Socioeconomic factors and financial strain

‘*Growing up in Ireland survey*’ calculated the index for LBW arising from PTB and intrauterine growth restriction (IUGR) and an association with parental education and environmental conditions was observed.^52^ The UK Bradford study found that a new variable of interest, financial strain, was associated with a significant increase in PTB.^53^ Direct and timely financial assistance offered by the Irish Government during the lockdown to businesses, self-employed and employees who were temporarily laid off, possibly avoided financial strain on the pregnant families.

#### 5. Maternal nutrition, opportunities for wellbeing and exercise

Observational data suggest the influence of maternal under nutrition in spontaneous PTB, which support a role for optimal maternal pre-pregnancy and in-pregnancy nutritional status in determining gestational length.^6^ A low maternal body-mass index (BMI) was associated with spontaneous PTB.^7^ Compared to no exercise during pregnancy, those taking appropriate leisurely exercise lowered the risk of PTB and placental weight gain could partially mediate the association between exercise during pregnancy and PTB.^54^ Irish lockdown allowed for outdoor exercise within a two kilometre radius of home and Government initiatives promoted online wellbeing and exercise choices through the ‘Healthy Ireland’ platform.^55^

#### 6. Reduction of chances for infections during pregnancy

Susceptibility to infectious diseases is often modified during pregnancy. Alterations in immunity to allow for the foetal allograft to implant and thrive combined with the anatomical and physiological modifications underlie these susceptibilities.^56^ It has been shown that relationship satisfaction reduces infectious diseases in pregnancy.^57^ Behavioural changes promoted during and prior to the lockdown, including social distancing, enhanced hand hygiene and use of face masks potentially reduced the chances of common viral infections during pregnancy. In addition, the closure of crèche, day-care centres, child minding facilities and play schools that normally ‘bring home’ common infections further reduced the potential for infective agent exposure.^58^ Adoption of WFH policies by pregnant women and their partners may have further reduced exposure to the ‘microbial world’ of the adult population. Consequently, there may have been a reduction in the likelihood of Influenza, parvovirus B19 and congenital cytomegalovirus (CMV) that are more significant infections during pregnancy, with acknowledged associations with PTB. ^58,59,60,61^

#### 7 Daily commuting and road traffic incidents

Irish national lockdown has brought down overall vehicular traffic, reduced the commuting to and from work locations, avoided early start and late return to home, attenuated traffic-related stresses and improved atmospheric air pollution. Arguably, the potential for crashes involving pregnant women should be low as well. After a single crash, pregnant drivers had increased rates of PPROM and PTB.^62,63^ Evaluation of potential adverse foetal outcome using a 26 weeks pregnant woman manikin has previously demonstrated significant harm using common accident scenarios.^64^ An observational study reported increased PTB following air travel, however large multi-centric studies would be warranted before drawing conclusions and the contribution to our study cohort would be low based on population characteristics.^65^

#### 8. Domestic abuse or intimate partner violence

During a lockdown or similar measures, depending on the socioeconomic factors and population characteristics, there is potential for upward or downward trends in domestic abuse or Intimate partner violence (IPV). The Irish Government launched proactive advertisement campaigns early in the lockdown period to educate and encourage measures against the potential for IPV. IPV is an important public health problem and an association between IPV and PTB has already been demonstrated. Prevalence of IPV was 14.9% in an Australian study and the main precipitating cause of PTB was antepartum haemorrhage.^66^ Psychological abuse by partner was associated with increased risk PTB in one South Indian study.^67^

#### 9. Cigarette smoke, coffee, alcohol, prescription drugs and street drugs

Approximately 11% of Irish women smoke during pregnancy and 28% of those who smoke while pregnant had SGA infants compared to non-smokers at 13%.^68^ Both active and passive (second-hand or environmental) tobacco smoking during pregnancy is associated with risk of SGA and PTB.^69,70^ PTB has been reported for women with drug dependence, cocaine and poly-substance being at the highest risk.^71^

Studies evaluating alcohol consumption during pregnancy are often overshadowed by bias attributable to unmeasured confounders and varying or no impact on PTB rates has been reported.^72^ Meta-analysis showed that high consumption of coffee during pregnancy is associated with low birth weight (LBW) and PTB.^73^ On the other hand certain herbal preparations claim myorelaxant, anti-inflammatory and immunomodulatory properties and were reported as useful in preventing PTB associated with inflammation and infection.^11^

While figures relating to cigarette smoking, alcohol consumption and coffee intake in pregnant women during the Irish lockdown are not yet available, it is hypothesized that with enhanced surveillance by enforcement agencies during this time, the availability, opportunities and the distribution for the illicit drug trade may have been reduced.

#### 10. Optimising maternal immunizations

Maternal immunization schedules are increasingly coming under the spotlight as part of the ‘life-course’ immunization programmes for the role that they play in improving maternal, foetal, and neonatal health.^74^ Even though not primarily targeted to reduce PTB, influenza vaccination during pregnancy indirectly reduces PTB through the reduction of maternal morbidity.^74^ Perhaps the lockdown afforded pregnant women the opportunity to optimize preventive approaches including immunizations; however ease of access to services apparently was not uniform.

### Learning from the lockdown and future strategies to reduce PTB

Only three VLBW and no ELBW infant admission to the only neonatal intensive care unit (NICU) of one of the health regions of Ireland from 1^st^ January to 30^th^ April of 2020, resulted in a reduction to zero of morbidity metrics primarily linked to extreme prematurity such as necrotising enterocolitis (NEC), retinopathy of prematurity (ROP) and severe forms (grade 3 and 4) of intraventricular haemorrhage (IVH).

In the absence of significant alterations to antenatal or perinatal hospital-based care pathways, our observed reduction of PTB among the VLBW infants points to the lockdown induced behavioural and socio-environmental modifiers as the catalysts for the reported change. We recommend varied and broadened preventive approaches based on sociodemography, nutrition, lifestyle, and underlying individual genetic and epigenetic variations.^75^ This finding of clinically meaningful reduction of PTB through non-clinical, socially rooted alterations in maternal behaviour and lifestyle along with an enabling environment, challenges the widely held assumption that the agents for prevention of PTB are hospital-based. These observations, if replicated from other regions of the world during the pandemic with varying levels of socio-environmental restrictions triggered by COVID-19, could offer novel perspectives and promising insights facilitating the analysis of yet under-appreciated phenotype of PTB. Statistical modelling approaches and big-data analysis principles would also be critical in this journey to reduce PTB.^16^

Standardized prenatal care and timely perinatal interventions could inherently lead to medically induced PTB, while rendering the desired reduction of foetal and maternal morbidity and stillbirths.^76,77^ Medicalization of the physiological process of pregnancy is often criticized for excessive monitoring and interventions, most notably in health systems rooted on fee payment at point-of-care, private health insurance or reimbursements.^78,79^ Based on our observations and reflecting on the changing demographics of pregnancy and childbirth, a contextual reconceptualization of antenatal care integrating the domains of health, behaviour, society and environment would appear more effective in reducing the VLBW and ELBW rates.

Our observed new PTB trend, if an outcome of the COVID-19 enforced socio-environmental and behavioural changes, suggest it is reasonable to postulate, a) the low numbers from early March onwards was influenced by the effects of the pre-lockdown period of extra public health vigilance that commenced in mid-February, b) effects are immediate, and c) the effects of lockdown will be seen in coming months and sustainable until such time as normality influencers are operating again. However, post-lockdown deterioration in socio-environmental factors or a ‘baby boom’ in late 2020 could increase the PTB rates.

### Limitations, Interpretations and Generalisability

The following limitations are acknowledged: 1. Inherent reservations posed by retrospective nature of the birth cohort data spanning over two decades; 2. Even though the vast majority of the VLBW infants would be premature, rarely severe intrauterine growth restriction (IUGR) at term could be included. However, such an inclusion consistently over two decades should reduce the bias and the weight-based inclusion criteria would allow comparison to national data by CSO, NPEC and internationally through VON; 3. Inclusion of January 2020, when there was no official lockdown or enhanced pre-lockdown public health measures. This was required to make comparison with the two decades of trends including the first four months; 4. Completion of the study prior to the official finish of lockdown was to facilitate ease of comparison against the coded historical monthly data, timely data completion and analysis; 5. ELBW category could only be analysed with limitations considering the small number of births on a monthly basis; 6. We caution the ‘no abortion policy’ that Ireland followed till late 2018, when making international comparisons; 7. The lockdown could have deferred what should have been medically offered early during pregnancy as well, thus arguably postponing the gestational age of intervention. Thus it could be viewed that the potential for reduced monitoring opportunities or early foeto-maternal interventions during the lockdown could have ‘shifted the band’ from VLBW to LBW infants; 9. Our observations are suggestive of potential associations of the socio-environmental and behavioural modifiers with reduced PTB and the study is not designed to evaluate causality. Moreover, our dataset from a regional sample in Ireland is relatively small and thus not strictly reflecting every society’s behavioural and socio-environmental response to the COVID-19 lockdown.

## Conclusion

The Irish national lockdown in response to SARS-CoV-2 virus (COVID-19) pandemic and the cumulative effects of socio-environmental variables such as maternal behavioural modifications, opportunities to work-from-home, potential reduction in work related stresses, possible alleviation of physical strain related to work and commuting, optimal opportunities for rest and sleep, likely increase in partner presence and support at home, reduced exposure to infections, improved opportunities for nutritional support and exercise as well as the positive alterations in environment and air pollution, all would have possibly resulted in a reduction in PTB involving ELBW and VLBW infants. In the context of achieving sustainable development goals (SDG) and to reduce the under-five mortality globally, prematurity rate would be the most important ‘*curve to bend*’.^80^ Further research is needed to enhance our knowledge regarding the complex ways in which environmental, social, behavioural and biological factors interact and modify PTB, an important perinatal event of global significance. We recommend WHO and national policy makers reflect on this positive outcome of the COVID-19 pandemic, the insight that it has provided, and seize the opportunity to recommend further studies from geographically diverse regions to evaluate these implicated interdependent behavioural and socio-environmental modifiers of PTB.

**Word count of manuscript:** 4637 (limit 5000)

## Data Availability

All the relevant data pertaining to the study have been included in the manuscript. No other online material or repositories is linked to this manuscript.

## Acknowledgements

Authors wish to acknowledge Thomas Stack, Gerard Burke, Con Sreenan, Niazy Al-Assaf, Margo Dunworth, Deirdre O’Connell, Marie Carroll, Collette Quinn, Irene Byrne, Leona Blackwell and Eamon Leahy for various stages of neonatal and perinatal activity tabulation and verification over the two decades at UMHL. Athanasios Mantas is acknowledged for the information related to the termination of pregnancy (TOP) and Grace McCormack for collating the labour ward activity at UMHL. John Slevin and Rizwan Khan (obstetric and neonatal professionals) as well as Sharon O’Brien (parent of a VLBW infant, as part of PPI) are appreciated for their independent scrutiny and verification of primary VLBW and ELBW data for 2020. Patrick Dillon and Joanne O’Connor are acknowledged for the research governance and ethics approvals. National Perinatal Epidemiology Centre (NPEC) is acknowledged for the recent Vermont Oxford Network (VON) Ireland information and the Central Statistics Office (CSO) of Ireland for the National data. Obstetric, midwifery, nursing and neonatal professionals of UMHL who otherwise supported the project through their clinical input and assistance are gracefully acknowledged.

## Footnotes

### Contributorship statement

RKP conceptualised and designed the study, extracted the ELBW and VLBW data, drafted the initial draft, reviewed and revised the manuscript. ER collected the neonatal activity data and contributed to the nursing and midwifery sections. HP developed the data analysis instruments, conducted the statistical analysis and reviewed and revised the relevant sections. MI contributed to the obstetric data, verified the termination of pregnancy register and reviewed the relevant sections. MD contributed as the patient and public representative, contributed to the relevant sections and developed the search strings. NO co-supervised the research and critically reviewed the infection and microbiology sections. DM co-supervised the research, reviewed and edited the relevant sections. CD coordinated and supervised the research, advised on methodology and manuscript structure and critically reviewed the manuscript for important intellectual content. All authors approved the final manuscript as submitted and agree to be accountable for the work. The corresponding author attests that all listed authors meet authorship criteria and that no others meeting the criteria have been omitted.

### Ethical approval

Research Ethics Committee of University Hospital Limerick, Ireland, approved the study.

### Data sharing

All relevant data included in the manuscript. No additional data available.

The corresponding author affirms that the manuscript is an honest, accurate, and transparent account of the study being reported; that no important aspects of the study have been omitted; and that any discrepancies from the study as planned have been explained.

### Dissemination to participants and related patient and public communities

There are no individual study participants to disseminate the results to. Neonatal patients advocacy charity (INHA) involved in the study will be sent a copy of the article when published. The authors intend to disseminate the study findings through INHA website and other media so that the results will be available for the wider patient and public communities.

---

**Supplementary Table 1:** Poisson Regression Analysis of VLBW counts Jan - April at UMHL

| Time Period | Births | VLBW | Rate (Wald 95% CI) | Rate Ratio (95% CI) | p-value |
| --- | --- | --- | --- | --- | --- |
| **2001 - 2019** | 29324 | 240 | 8.18 (7.21, 9.29) | 3.77 (1.21, 11.75) | 0.022 |
| **2020** | 1381 | 3 | 2.17 (0.70, 6.74) |  |  |

**Supplementary Table 2:** Annual VLBW and ELBW counts and rates per 1000 live births at UMHL and nationally.

|  | Time Period | Births | VLBW | Rate (Wald 95% CI) | ELBW | Rate (Wald 95% CI) |
| --- | --- | --- | --- | --- | --- | --- |
| **UMHL** | 2001 - 2019 | 89029 | 749 | 8.41 (7.83, 9.04) | 255 | 2.86 (2.53, 3.24) |
| **National CSO** | 2011 - 2017 | 473157 | 4225 | 8.93 (8.66, 9.20) | 1615 | 3.41 (3.25, 3.58) |
| **National VON** | 2014 - 2017 | 258496 | 2343 | 9.37 (9.01, 9.75) | 935 | 3.62 (3.39, 3.86) |

